# Association between gut microbiome composition and symptom self-report in trauma exposed OEF/OIF/OND Veterans

**DOI:** 10.1101/2023.11.01.23297931

**Authors:** Y. Irina Li, Kathleen Pagulayan, Holly Rau, Rebecca Hendrickson, Abigail G. Schindler

## Abstract

**Objective:** Iraq and Afghanistan war-era (OEF/OIF/OND) Veterans are at elevated risk for physical injuries and psychiatric illnesses, in particular comorbid mild traumatic brain injury (mTBI), posttraumatic stress disorder (PTSD), and chronic pain. The gut microbiome has been implicated in modulation of critical processes such as digestion, immune system functioning, and stress responsivity, and may be an important factor in understanding physical and mental health outcomes following deployment and trauma exposure, yet minimal research to date has sought to characterize gut microbiome composition in this population.

**Methods:** 26 male OEF/OIF/OND Veterans aged 18 to 65 who previously completed a VA Comprehensive TBI Evaluation were enrolled in this study. Participants completed self-report measures of PTSD symptom severity, pain intensity and interference, fatigue, cognitive symptoms, substance use, and sleep quality. Participants submitted fecal samples, and metagenomic sequencing was used to calculate alpha- and beta-diversity and taxonomic microbial composition. Associations between microbiome data and clinical variables was then examined.

**Results:** Alpha and beta diversity measures were not significantly correlated with clinical outcomes. Fatigue, post-concussive symptoms, executive function symptoms, and cannabis use were associated with differences in gut microbial composition, specifically Verrucomicrobiota.

**Conclusion:** This exploratory study demonstrated that altered gut microbiome composition is associated with psychiatric and cognitive symptoms in OEF/OIF/OND Veterans and highlights a potential new therapeutic target of interest. Future research is needed to examine whether probiotic treatment is effective for reducing symptoms common in this clinical population.

## Background

The negative impact on physical and mental health following deployment to war zones among U.S. military personnel and Veterans has been well documented in the empirical literature (Hoge et al., 2004; Milliken, Auchterlonie, & Hoge, 2007; Spelman, Hunt, Seal, & Burgo-Black, 2012; Vogt, Pless, King, & King, 2005). Data from the Millennium Cohort Study—a prospective longitudinal study by the departments of Veterans’ Affairs (VA) and Department of Defense—showed that deployment is associated with increased cardiovascular risk, smoking initiation, chronic pain, and greater medical illness burden (Brown, 2010; Cohen, Marmar, Ren, Bertenthal, & Seal, 2009; Frayne et al., 2011; Gironda, Clark, & Walker, 2005; Granado et al., 2009). Likewise, in a large-scale review, Pietrzak and colleagues found increased rates of post-traumatic stress disorder (PTSD), depression, and anxiety post-deployment (Pietrzak, Pullman, Cotea, & Nasveld, 2012). Despite greater understanding in recent years of the negative impact of deployment and combat exposure on health outcomes, the full breadth of underlying mechanisms are still being uncovered and effective treatment options are limited.

### The Polytrauma Clinical Triad in Returning Veterans

Service Members involved in Operations Enduring Freedom, Iraqi Freedom, and New Dawn (OEF/OIF/OND) experience greatly elevated rates of co-occurring mild traumatic brain injury (mTBI), PTSD, and pain, commonly referred to as the “polytrauma clinical triad” (Lew et al., 2009). Increasing evidence indicates that the presence of some combination of the polytrauma triad make individuals particularly vulnerable to additional medical and psychiatric comorbidities and lead to greater overall chronic disease burden (Hendrickson, Schindler, & Pagulayan, 2018; Pugh et al., 2014; Zouris, Wade, & Magno, 2008). For example, the combination of deployment-related mTBI and PTSD is associated with greater PTSD and depressive symptoms, pain interference, and poorer sleep quality (Martindale et al., 2020). Newly emerging evidence also suggests that mTBI sustained in the context of combat leads to poorer outcomes and greater disability relative to those service members who did not sustain a TBI, even a decade later (Mac Donald, Barber, Johnson, Patterson, & Temkin, 2022). Thus, greater understanding of multifactorial influences on long-term outcomes in this unique and complex population is critical for the development of effective interventions to better improve outcomes.

### Potential Role of the Gut Microbiome in Modulating Health Outcomes

The gastrointestional (GI) system may be particularly vulnerable to the effects of blast exposures, as well other deployment related factors. Indeed, there is growing recognition that deployment-related factors can impact GI health (Maguen, Madden, Cohen, Bertenthal, & Seal, 2014; Vanderploeg et al., 2012). Critically, an accumulating body of literature suggests that a positive link exists between GI distress and mental health disorders (Bener, Ghuloum, Al-Hamaq, & Dafeeah, 2012; Spiegel et al., 2011), which may in part contribute to the complex clinical picture of overlapping adverse physical and psychological outcomes in returning Service Members and Veterans.

The association between psychological and GI distress has led to increasing attention around the role of the microbiome in the modulation of physical and mental health symptoms. The human microbiota, referring to all microorganisms in and on the human body, is made up of bacteria, archaea, fungi, protozoa, and bacteriophages (Dave, Higgins, Middha, & Rioux, 2012). The majority of bacteria that make up the human microbiome reside in the gut and can be impacted by a wide range of factors, including genetics, environment, early childhood experiences, physical illness, physical activities, and medications (Goodrich et al., 2016; Jackson et al., 2018; Maier & Typas, 2017; Munyaka, Khafipour, & Ghia, 2014). In turn, the gut microbiome plays a crucial role in modulating numerous functions important for health and survival, including digestion and immune system functioning (D’Argenio & Salvatore, 2015). In addition, the gut microbiome is thought to play an important role in bidirectional communication between the gut and brain (the microbiota-gut-brain axis, or MGBA), particularly in the context physical and psychological trauma (Montiel-Castro, González-Cervantes, Bravo-Ruiseco, & Pacheco-López, 2013; Overman, Rivier, & Moeser, 2012; Zhu, Grandhi, Patterson, & Nicholson, 2018).

While the literature is still emerging, studies suggest that the gut microbiome is implicated in bidirectional signaling of affective processes. Modification of the gut microbiome via colonization of donor gut bacteria from mice with enhanced anxiety phenotype has been shown to elicit anxiety-like behaviors in non-anxious mice (Bercik et al., 2011). Alteration of the gut microbiome via administration of beneficial bacteria such as *Lactobacilius rhamnosus* or *Bifidobacterium longum* has led to decreased anxiety and depressive behaviors in rodent models (Bercik et al., 2011; Bravo et al., 2011). In humans, probiotic administration has been shown to reduce anxiety and depression symptoms (Pirbaglou et al., 2016), supporting MGBA as a potentially important modulatory link between physical and mental health.

Stress exposure may impact gut microbial composition via the release of stress hormones or increased gut permeability, altering the microbiota habitat (Montiel-Castro et al., 2013; Overman et al., 2012). Gut microbiome composition can be influenced by both distal stressors such as early life experiences and proximal stress such as blast and TBI (Leclercq, Forsythe, & Bienenstock, 2016; O’Mahony et al., 2009; Wang, Zhu, Hou, & Hou, 2021). Gut microbiome has been shown to play a role in the modulation of systems involved in affective signaling including the hypothalamic-pituitary-adrenal axis (Sudo et al., 2004), which may increase susceptibility to emotion dysregulation and subsequent risk for the development of psychiatric conditions following exposure to trauma.

Taken together, evidence suggests that the gut microbiome may be an important factor in understanding the trajectory of physical and mental health outcomes of military personnel following deployment, particularly in the context of the polytrauma clinical triad. The limited available literature on gut microbiome in Veterans has predominantly focused on broadly characterizing Veterans as a whole, or in cohorts based on involvement in foreign conflicts or disease status (Bajaj et al., 2019; Janulewicz et al., 2019; Keating et al., 2019; Stanislawski et al., 2021), and has not focused on psychological or mental health outcomes nor specifically on combat Veterans of the OEF/OIF/OND conflicts. Therefore, enhancing our understanding of gut microbiota composition in this unique population is critical in elucidating potential pathogenic processes involved in the polytrauma clinical triad and potentially informing future therapeutic targets. The goals of the present study were to examine (a) the gut microbiome profile in a sample of combat deployed OEF/OIF/OND Veteran, and (b) potential associations between gut microbial composition and physical and mental health symptoms.

## Methods

### Study Procedures

Prospective study participants were identified via a review of the VA Computerized Medical Record System for Veterans who completed a Comprehensive TBI Evaluation (CTBIE) at a large metropolitan Veterans Health Administration (VHA) facility between 2019 and 2021. Individuals were eligible for the study if they were an OEF/OIF/OND Veteran between the ages of 18 and 65 with a remote history of a blast-related mTBI or no history of TBI in adulthood, and were able to read, speak, and comprehend English. As this was an exploratory study with a relatively small sample, only male Veterans were eligible given that the majority of OEF/OIF/OND Veterans are male. Exclusion criteria were a) mTBI within the past 3 months, b) moderate or severe TBI diagnosis, c) history of serious mental illness (e.g. schizophrenia, schizoaffective disorder, or bipolar disorder), d) current (defined as past 3 months) substance use disorder diagnosis, e) current neurologic disorder that could impact cognitive functioning, f) high risk of suicide, or g) failure of two or more items on a brief 6-item cognitive screener. The study was approved by the local Institutional Review Board.

After providing informed consent, study participants were asked to complete an online survey and provide a fecal sample collected at home using the OMNIgene GUT self-collection kit. Fecal samples were returned to the research team via overnight mail delivery and then sent to Diversigen for sample processing and bioinformatic analysis. Online surveys were completed within 3 days of sample collection included questions about demographic information, medical and psychiatric history, and current use of medications and over-the-counter supplements.

### Clinical Symptom Measures

TBI history was gathered from Veterans’ electronic health records via the CTBIE, a VA-mandated semi-structured interview that is completed by a physician as part of clinical care. PTSD symptom severity was assessed with the PTSD Checklist for DSM-5 (Weathers et al., 2013). Depression symptom severity was assessed with the Patient Health Questionnaire (PHQ-9; Kroenke et al., 2001). Fatigue was measured with the Patient-Reported Outcomes Measurement Information System (PROMIS; Cella et al., 2010) Fatigue Short Form 6a. Pain severity was measured using the Pain Intensity Short Form 3a (Revicki et al., 2009) while interference of pain on daily activities was assessed with PROMIS-Pain Interference Short Form 8a (Amtmann et al., 2010). The Neurobehavioral Symptoms Inventory (NSI; Cicerone & Kalmar, 1995) was used to assess current (past two weeks) post-concussive symptoms. Current cognitive symptoms were assessed with the Traumatic Brain Injury-Quality of Life Measurement System (TBI-QOL) Attention and Concentration, Executive Functioning, and Learning and Memory scales (Tulsky & Kisala, 2019; Tulsky et al., 2016). Sleep quality was assessed with the Pittsburgh Sleep Quality Index (PSQI; Buysse, Reynolds, Monk, Berman, & Kupfer, 1989). Current substance use was assessed with the Alcohol Use Identification Test (AUDIT; Saunders et al., 1993), Cannabis Use Disorder Identification Test-Revised (CUDIT-R; Adamson et al., 2010), and Drug Abuse Screening Test (DAST; Skinner, 1982). For all participant-reported symptom measures with exception of the TBI-QOL, higher scores reflect greater symptom severity.

### Fecal sample collection

The OMNIgene GUT self-collection kit was used by study participants at home to collect stool specimen. This system allows for rapid homogenization and stabilization of microbial DNA at point-of-collection, eliminating cold chain requirement (sample storage at ambient room temperature for up to 60 days). Briefly, each participant was instructed to 1) deposit stool into a collection bowl suspended over a toilet, 2) transfer an aliquot of stool using the provided scoop into the collection tube (contains homogenization bead and solution to stabilize microbial DNA), 3) shake sealed tube for 30 seconds, and 4) return sample in pre-addressed mailing envelope to the VA study site.

### Fecal sample processing

Fecal samples were mailed in bulk to Diversigen for DNA extraction and metagenomic analysis using their BoosterShot Shallow Shotgun Sequencing. In brief, DNA sequences were aligned to a curated database containing all representative genomes in RefSeq for bacteria. Alignments were made at 97% identity against all reference genomes. Every input sequence was compared to every reference sequence in the Diversigen Venti database using fully gapped alignment with BURST. Ties were broken by minimizing the overall number of unique Operational Taxonomic Units (OTUs). For taxonomy assignment, each input sequence was assigned the lowest common ancestor that was consistent across at least 80% of all reference sequences tied for best hit. Samples with fewer than 10,000 sequences were discarded (no samples in the current study were discarded). OTUs accounting for less than one millionth of all strain-level markers and those with less than 0.01% of their unique genome regions covered (and < 0.1% of the whole genome) at the species level were discarded. The number of counts for each OTU was normalized to the OTU’s genome length.

### Data analysis

Centered log ratio transformations were were conducted on phylum-level compositional data to account for non-normality in distribution of microbiome data. We then conducted non-parametric Spearman correlations of gut microbiome parameters with clinical outcome variables. An additional analysis examined rank-biserial correlations between gut microbiome parameters and self-report medication use. We elected to specifically examine medications with strong documented effects on the gut microbiome, which in our sample were medications with serotonergic action (selective serotonin reuptake inhibitors, selective serotonin receptor agonists, serotonin and norepinephrine reuptake inhibitors, tricyclic antidepressants), angiotensin-converting enzyme (ACE) inhibitors, alpha and beta blockers, and Vitamin D (Vich Vila et al., 2020; Weersma, Zhernakova, & Fu, 2020). Self-reported health conditions were categorized into mental health conditions (depression, PTSD, generalized anxiety disorder, and ADHD), neurological disorders (migraine/headaches, stroke or aneurysm, brain tumor, brain surgery, epilepsy or seizures, multiple sclerosis, Parkinson’s Disease, and other neurological illness), gastrointestinal disorders (GERD, gluten intolerance, Celiac Disease, lactose intolerance, IBS, IBD, and other gastrointestional problem), endocrine disorder (liver disease, kidney disease, Hepatitis C, thyroid disease, low testosterone, growth hormone deficiency, and other endocrine condition), sleep disorders (insomnia and sleep apnea), and other health conditions (hypertension, hypercholesterolemia, hyperlipidemia, cardiovascular disease, heart attack, diabetes, cancer, asthma, chronic obstructive pulmonary disease, B12 deficiency, musculoskeletal pain, arthritis, gout, and lupus). Correlational analyses were conducted using SPSS 26.0.0 with correction for multiple comparisons using the Benjamini-Hochberg False Discovery Rate method.

Alpha diversity indices assessed were observed species richness, Chao1, and Shannon, measuring diversity within samples. Beta diversity (measuring dissimilarity between samples) was evaluated using Bray Curtis dissimilarity matrix and visualized using principal coordinate analysis (PCoA). Biplots were used to visualize associations between participant samples and clinical self-report variables. Biplot analysis were conducted in R (2022.07.1).

## Results

### Participant demographics and polytrauma clinical triad assignment

A total of 26 participants (mean age = 41.9 years, 50% White, 12% Black or African American, 4% Asian or Asian American, 4% American Indian or Alaskan Native, 8% Native Hawaiian or Pacific Islander, 12% Mixed, and 12% Other; 15% Hispanic ethnicity) were enrolled in the study. Table 1 summarizes demographic data and clinical symptom measures of study participants. Approximately half (57%) of participants had three or more deployments, and spent in total one or more years on deployment. Figure 1 shows the proportion participants with polytrauma clinical triad symptoms in this sample. Approximately 62% of Veterans had a history of at least one mTBI during deployment. Approximately 81% of the participants had a self-reported lifetime diagnosis of PTSD, and approximately 54% met the clinical cut-off for probable PTSD diagnosis on the PCL-5 at the time of the study (i.e., based on a clinical cutoff of 33; (Bovin et al., 2016). Clinically significant pain intensity was reported by 69% of participants, and 73% reported clinically significant pain interference (i.e., based on T-scores higher than one standard deviation above community norms; (Sager, Wachen, Naik, & Moye, 2020). Only two participants did not endorse clinically significant PTSD symptoms, pain symptoms, or history of mTBI. One or more gastrointestinal disorder diagnosis was reported by 42% of Veterans.

**Figure 1.**
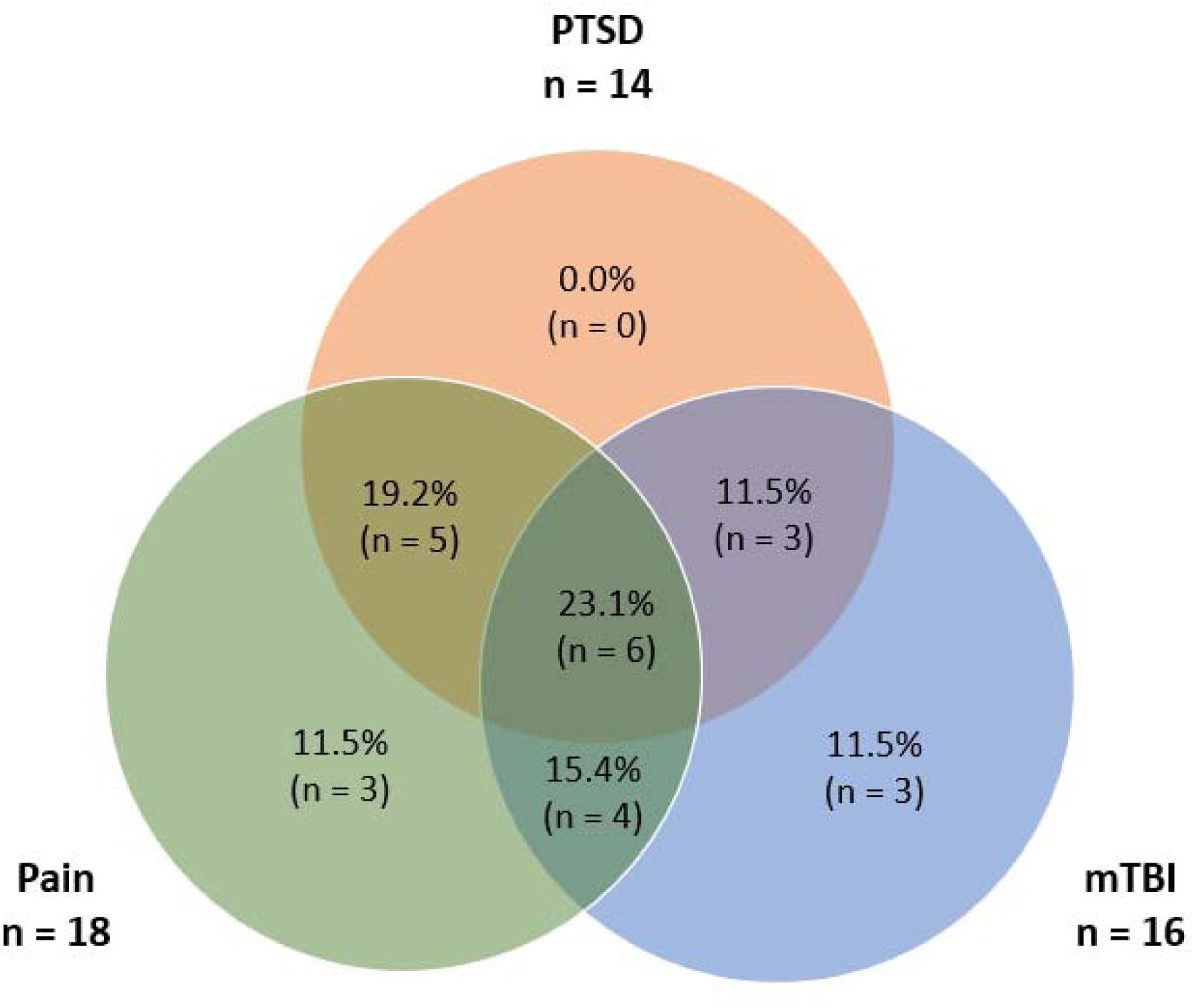
The Polytrauma Clinical Triad: Distribution of study participants with clinically significant post-traumatic stress disorder (PTSD) symptoms, chronic pain interference, and mild traumatic brain injury (mTBI).

**Table 1.** Characteristics of Study Participants (N = 26)

| **Demographic Variables** |  |
| --- | --- |
| Age (M (SD)) | 41.9 (7.2) |
| Race (n) |  |
| White | 13 |
| Black | 3 |
| Asian | 1 |
| American Indian or Alaskan Native | 1 |
| Native Hawaiian or Pacific Islander | 2 |
| Mixed Race | 3 |
| Other | 3 |
| Hispanic ethnicity (n) | 4 |
| Education level (n) |  |
| High school graduate or GED | 4 |
| Some college | 6 |
| Associate's degree | 3 |
| Bachelor's degree | 7 |
| Graduate degree | 6 |
| **Medications with Influences on the Gut Microbiome (n)** |  |
| Serotonergic system acting | 10 |
| ACE inhibitors | 5 |
| Alpha or beta blockers | 7 |
| Vitamin D | 12 |
| **Clinical Variables** | **M (SD)** |
| Number of self-reported lifetime health conditions |  |
| Mental health disorders | 2.2 (1.3) |
| Neurological disorders | 0.8 (0.6) |
| Gastrointestinal disorders | 0.6 (0.9) |
| Endocrine disorders | 0.2 (0.4) |
| Sleep disorders | 1.0 (0.8) |
| Other health disorders | 2.4 (1.4) |
| PROMIS |  |
| Pain Intensity | 25.3 (8.6) |
| Pain Interference | 9.0 (2.0) |
| Fatigue | 19.8 (6.3) |
| PSQI | 11.8 (3.7) |
| PHQ-9 | 12.5 (7.5) |
| PCL-5 | 35.3 (19.5) |
| NSI | 33.3 (17.0) |
| TBI-QOL |  |
| Attention/Concentration | 16.9 (6.4) |
| Executive Functioning | 34.0 (10.2) |
| Learning/Memory | 18.6 (5.9) |
| AUDIT | 3.5 (6.1) |
| CUDIT-R | 2.4 (3.4) |
| DAST | 1.1 (1.5) |

Note. Mental health disorders = depression, PTSD, generalized anxiety disorder, and ADHD. Neurological disorders = migraine/headaches, stroke or aneurysm, brain tumor, brain surgery, epilepsy or seizures, multiple sclerosis, Parkinson's Disease, and other neurological illness. Gastrointestinal disorders = GERD, gluten intolerance, Celiac Disease, lactose intolerance, IBS, IBD, and other gastrointestinal problem. Endocrine disorders = liver disease, kidney disease, Hepatitis C, thyroid disease, low testosterone, growth hormone deficiency, and other endocrine condition. Sleep disorders = insomnia and sleep apnea. Other health disorders = hypertension, hypercholesterolemia, hyperlipidemia, cardiovascular disease, heart attack, heart surgery, diabetes, cancer, asthma, chronic obstructive pulmonary disease, B12 deficiency, musculoskeletal pain, arthritis, gout, and lupus. PROMIS = Patient-Reported Outcomes Measurement Information System. PSQI = Pittsburgh Sleep Quality Index. PHQ-9 = Patient Health Questionnaire-9. PCL-5 = PTSD Checklist for DSM-5. NSI = Neurobehavioral Symptoms Inventory. TBI-QOL = Traumatic Brain Injury-Quality of Life. AUDIT = Alcohol Use Disorders Identification Test. CUDIT-R = Cannabis Use Disorders Identifications Test-Revised. DAST = Drug Abuse Screening Test.

### Correlation between gut microbiome and self-reported clinical symptoms

Table 2 shows a heat map of the correlations of gut microbiome parameters and polytrauma triad-related clinical outcome variables. In the analysis of alpha diversity measures, none were significantly correlated with any current clinical symptoms. While observed species number and Chao1 index were marginally negatively correlated with pain interference, fatigue, and sleep quality, results were not significant when adjusting for multiple comparisons. In the analysis of beta diversity measures, when correcting for multiple comparisons, no significant correlations were found (fatigue, sleep quality, and cannabis use were all marginally correlated prior to multiple comparison correction).

**Table 2.**
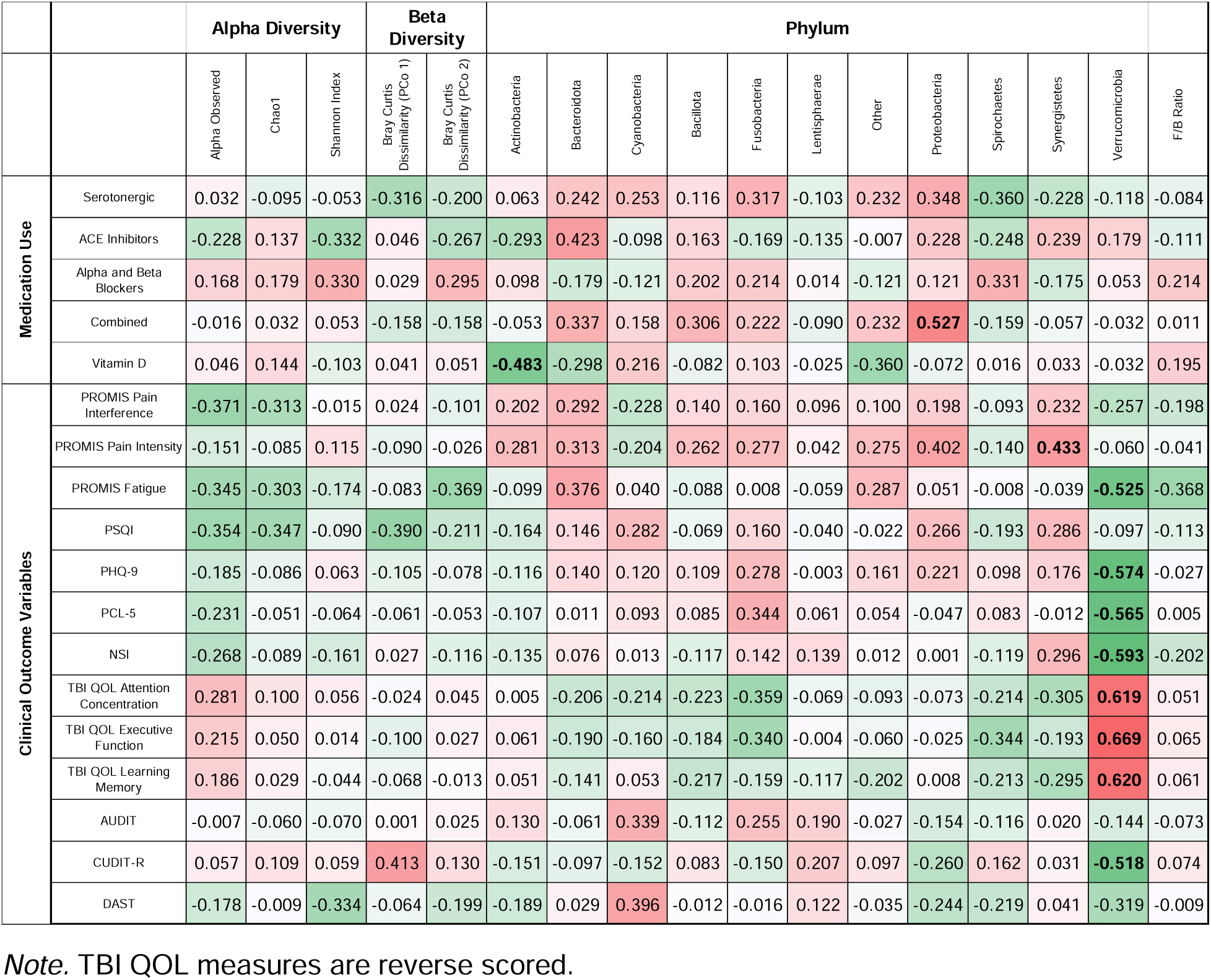
Heat map of non-parametric Spearman correlations between gut microbiome diversity metrics and clinical outcome measures. Bolded r-values values are significant at FDR 0.05.

At the phylum taxonomic level, abundance of Actinobacteria was significantly negatively correlated with use of vitamin D and Proteobacteria was significantly positively associated with combined medication use. Verrucomicrobiota showed the strongest associations with self-reported symptoms, with decreased abundance associated with greater fatigue, depressive symptoms, PTSD symptom severity, postconcussive symptoms, cannabis use, as well as difficulties with attention, executive function, and learning and memory.

### Biplot analysis of gut microbiome diversity

PCoA biplots were performed to further visualize and identify associations between participant fecal samples in relation to self-report measures (Figure 2a-e). A biplot displays a dimensionality reduction of samples (e.g., PCoA generated from OTUs using Bray Curtis) and variables of interest in self-report measures with respect to the same set of coordinates. Variables of interest are projected as vectors with length of vector indicating strength of association and direction indicating degree of difference between PCoA axes. Biplots are shown for a) phylum-level taxonomy; b) polytrauma clinical triad self-report; c) cognitive self-report; d) substance use self-report; and e) medication class use self-report. Among phylum-level taxonomy, Bacteroidota and Bacillota (Firmicutes) were the strongest drivers of differences in microbiome composition and were aligned with samples that were different in gut microbiota compositions. Among clinical outcome variables, fatigue and PCS symptoms were most strongly correlated with microbiome composition. Of the cognitive self-report measures, executive functioning symptoms were the most strongly associated with differences in microbiome composition. Alcohol use, ACE inhibitor, alpha- and beta-blockers, and serotonergic medications were also associated with differences in gut microbiome composition.

**Figure 2.**
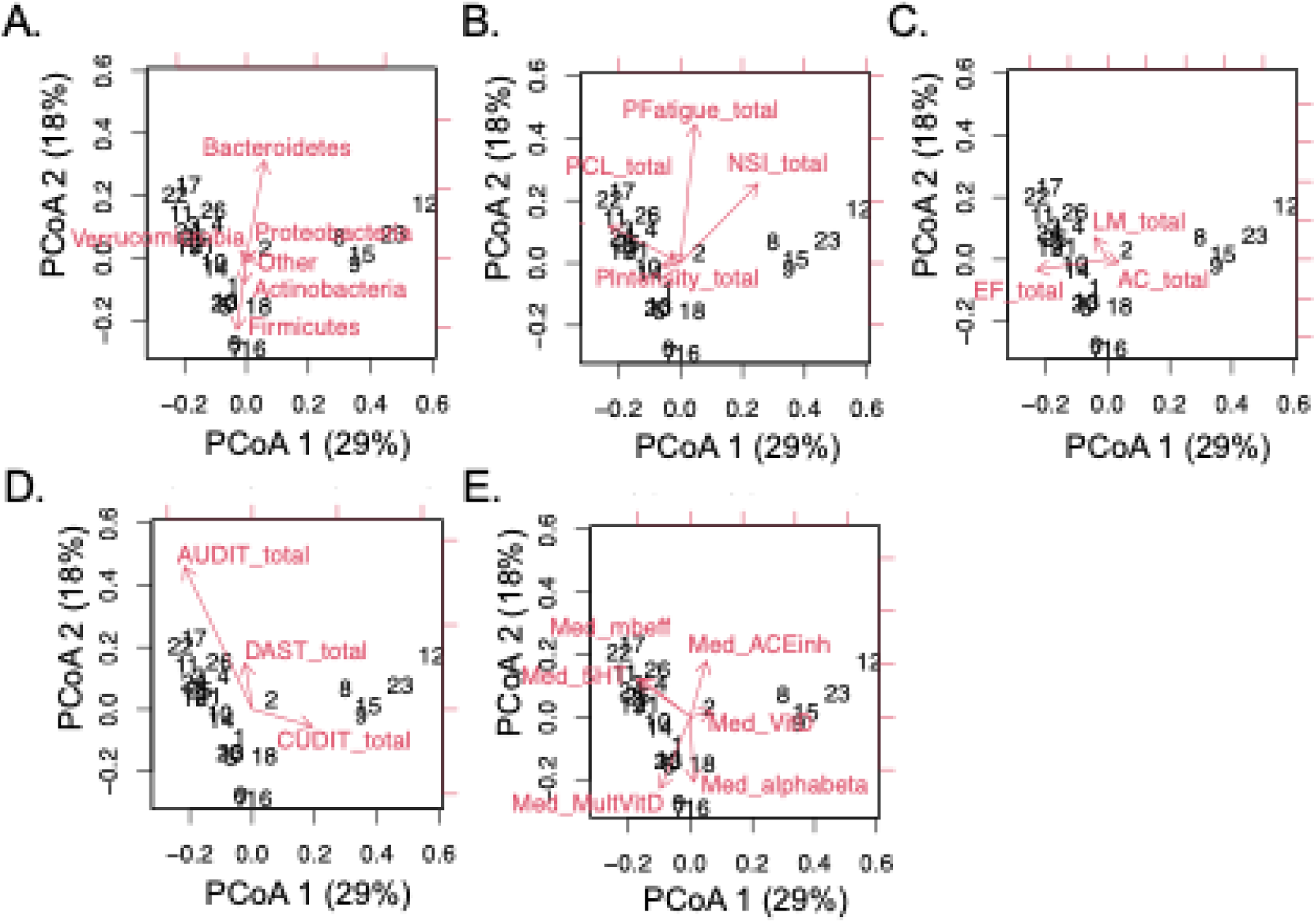
PCoA biplots displaying a dimensionality reduction of samples (e.g., PCoA generated from OTUs using Bray Curtis) and variables of interest in self-report measures with respect to the same set of coordinates. *Note.* Variables of interest are projected as vectors with length of vector indicating strength of association and direction indicating degree of difference between PCoA axes. A.=Phylum-level taxonomy; B.=Polytrauma clinical triad self-report; C.=Cognitive self-report; D.= Substance use self-report; E.=Medication class use self-report. PCoA: Princple Coordinate Analysis; LM: TBI-QOL learning and memory subscale; EF: TBI-QOL executive function subscale; AC: TBI-QOL attention and concentration subscale.

## Discussion

This exploratory study sought to characterize gut microbiome composition in a sample of OEF/OIF/OND Veterans who present with high rates of mTBI, PTSD, and pain. While the analyses did not reveal significant associations between measures of alpha or beta diversity and self-reported mental health or cognitive symptoms, a decreased abundance of the bacterial phylum Verrucomicrobia was strongly correlated with greater severity of PTSD and depressive symptoms, fatigue, and postconcussive symptoms. Verrucomicrobia has been positively linked with improved insulin sensitivity and glucose tolerance, and has been proposed to be protective for metabolic health (Niigiyimana et al., 2022; Zhang et al., 2023). Furthermore, our results in are line with a previous study in Veterans demonstrating a negative associated between Verrucomicrobia abundance and PTSD self-report (Hemmings et al., 2017). Likewise, in a rodent model of PTSD, Verrucomirobia exhibited both immediate and prolonged alteration in relative abundance following stress exposure (Gautam et al., 2018). Together, our results are consistent with previous reports, and raise the possibility that the prolonged stress associated with deployment (and repeated deployment for the majority of our sample) as well as chronic PTSD, may be associated with alterations in Verrucomicrobia abundance. Our findings provide preliminary support for altered gut microbiome composition in polytrauma clinic triad patients and highlight a potential new therapeutic target for OEF/OIF/OND with ongoing symptomology related to the polytrauma clinical triad.

Our results further suggest that fatigue may also be linked with differences in gut microbial community composition. Persistent fatigue is a frequent complaint in conditions such as chronic pain and IBD, and is a complex multifactorial clinical problem with a number of proposed pathways including central nervous system dysfunction, inflammation, dysregulation of sleep-wake mechanisms, and altered motivational and reward-processing functioning (Borren, van der Woude, & Ananthakrishnan, 2019; Van Damme, Becker, & Van der Linden, 2018). Interestingly, bacteriotherapy via transcolonoscopic infusion of non-pathogenic enteric bacteria has been shown to improve symptoms in patients with Chronic Fatigue Syndrome (Borody, Nowak, & Finlayson, 2012), suggesting the relationship between fatigue and gut microbiome may be bidirectional. Fatigue is also commonly reported in Veterans with mTBI, particularly in the presence of comorbid sleep disturbances, depression, and chronic pain (Mortera, Kinirons, Simantov, & Klingbeil, 2018; Rau et al., 2018). In the present study, both fatigue and *Bacteroidetes* were aligned with fecal samples with similar gut microbiome characteristics, suggesting that examination of potential associations between fatigue and *Bacteroidetes* may be important for understanding clinical outcomes in the polytrauma clinical triad population.

Postconcussive symptoms were also a factor in accounting for similarities in microbial community structure in this sample. Patients with TBI are at greater risk for gastrointestinal dysfunction and distress (Katzenberger, Ganetzky, & Wassarman, 2015); it has been suggested that downstream negative consequences of TBI, such as increased incidence of disease and disability, are rooted in part in dysregulation of the gut-brain axis following brain injury (Hanscom, Loane, & Shea-Donohue, 2021; Ross, Ball, Sullivan, & Caroff, 1989). Repetitive mTBI has been found to lead to changes in microbial composition in the rat jejunum, with mTBI rats exhibiting reductions in gut microbial diversity compared to controls (Matharu et al., 2019). It is important to note that the findings have been somewhat inconsistent, with another study examining a model of repetitive mTBI in male mice demonstrating only small and transient alterations in the gut microbiome (Angoa-Pérez et al., 2020). A systematic review of animal and human studies suggests that TBI leads to alterations in gut microbiome composition, although the authors noted limitations involving small number of studies and sample sizes (Pathare et al., 2020). While studies of uncomplicated mild TBI in civilian populations have generally found that the vast majority of individuals recover quickly and with a relatively short-term course of post-concussive symptoms (Carroll et al., 2004; Iverson, 2005), more recent literature has suggested there is significant heterogeneity in outcome trajectory following mTBI even in civilian populations, with a subgroup of individuals continuing to experience symptoms and associated functional impairment one year post-injury (Dikmen, Machamer, & Temkin, 2017). Together, this suggests that changes in gut microbiota composition resulting from neurotrauma could in turn contribute to further pathogenic processes such as dysregulation of immune functioning (Ochoa-Repáraz, Mielcarz, Begum-Haque, & Kasper, 2011; Zhu et al., 2018).

### Limitations

A number of important limitations to the present study exist. First, this was an exploratory study with a small sample size, which reduced our power to detect significant effects. In addition, our sample of OEF/OIF/OND Veteran had high comorbid rates of mTBI, PTSD, and pain, consistent with other studies in this population. While this was expected given that our aim was to broadly characterize the gut microbiome in this population, high symptom burden and use of medications that may affect the microbiome preclude drawing conclusions about directionality of effects. There was also variability in the clinical characteristics in our sample, including in length of mTBI history, PTSD symptom severity, and use of non-medication supplements, as well as length of deployment, all of which are likely associated with differences in gut microbiota composition. Lastly, relative abundance of gut microbiota also demonstrate natural oscillations linked to circadian rhythmicity and the light-dark cycle (Matenchuk, Mandhane, & Kozyrskyj, 2020). Variability in timing of fecal sample collection may make disentangling the effects of the light-dark cycle from observable differences in gut microbiota composition difficult.

### Conclusion

The limited available literature examining the relationship between the gut microbiome and physical and psychological outcomes in Veterans is mixed and requires further investigation. To our knowledge, this is the first study to examine the gut microbiome in a sample of OEF/OIF/OND Veterans with high rates of polytrauma triad symptoms. Our results demonstrate potential alterations in gut microbiota composition associated with increased psychiatric and cognitive symptoms in Veterans following deployment to combat zones. OEF/OIF/OND Veterans face increased rates of deployment-related stress exposures as well as alterations in environmental factors such as living conditions, sleep patterns, dietary intake and eating patterns, and local microbiomes, which may place them at unique risk for changes in gut microbiome composition and downstream pathogenic processes. The present study joins an expanding body of literature examining the relationship between gut microbiota and physical and mental health in Veterans. Additional research is warranted to determine whether treatments targeting the gut (e.g., probiotics) have a salutary effect on symptoms common this this clinical population.

## Data Availability

All data produced in the present study are available upon reasonable request to the authors

## DISCLAIMER

The views expressed in this scientific presentation are those of the author(s) and do not reflect the official policy or position of the U.S. government or Department of Veteran Affairs.

## DECLARATIONS

### Competing interests

The authors declare that the research was conducted in the absence of any commercial or financial relationships that could be construed as a potential conflict of interest.

### Funding

This work was supported by grants from the Department of Veteran Affairs (VA) Basic Laboratory Research and Development (BLR&D) Career Development Award 1IK2BX003258 (AGS), Department of Veteran Affairs VISN 20 MIRECC (AGS, KP, RH, HR), University of Washington Garvey Institute (AGS, KP, RH)

### Authors’ contributions

The work presented here was carried out in collaboration among all authors. YIL, KP, HR, RH, and AGS contributed to conception and design of the study. YIL, KP, HR, RH, and AGS collected and analyzed data. YIL and AGS wrote the first draft of the manuscript. All authors contributed to manuscript revision, read, and approved the final manuscript.

## Acknowledgements

We would like to thank Sean Meichiel for considerable technical assistance.

## Notes

### Competing Interest Statement

The authors have declared no competing interest.

### Author Declarations

The study was approved by the Veterans Affairs Puget Sound Health Care System Institutional Review Board.

